# Description of a University COVID-19 Outbreak and Interventions to Disrupt Transmission, Wisconsin, August – October 2020

**DOI:** 10.1101/2021.05.07.21256834

**Authors:** Dustin W. Currie, Gage K. Moreno, Miranda J. Delahoy, Ian W. Pray, Amanda Jovaag, Katarina M. Braun, Devlin Cole, Todd Shechter, Geroncio C. Fajardo, Carol Griggs, Brian S. Yandell, Steve Goldstein, Dena Bushman, Hannah E. Segaloff, G. Patrick Kelly, Collin Pitts, Christine Lee, Katarina M. Grande, Amanda Kita-Yarbro, Brittany Grogan, Sara Mader, Jake Baggott, Allen C. Bateman, Ryan P. Westergaard, Jacqueline E. Tate, Thomas C. Friedrich, Hannah L. Kirking, David H. O’Connor, Marie E. Killerby

**Author notes:** These authors contributed equally to this work (Dustin W. Currie, Gage K. Moreno). **Disclosures** The findings and conclusions in this report are those of the authors and do not necessarily represent the official position of the Centers for Disease Control and Prevention. Use of trade names is for identification only and does not imply endorsement by the Centers for Disease Control and Prevention. **Author Contributions:** Drafting Manuscript: DWC, GKM, MEK, **Reviewing and Editing Manuscript:** MJD, IWP, AJ, KMB, DC, TS, GCF, CG, BSY, SG, DB, HS, GPK, CP, CL, KMG, AK-Y, BG, SM, JB, ACB, RPW, JET, TCF, HLK, DHO, MEK, **Conceptualization and Data Collection Procedures:** DWC, GKM, IWP, KMB, KMG, AK-Y, BG, SM, RPW, JET, TCF, HLK, DHC, MEK, **Designing and Implementing Policies and Interventions:** DC, TS, CG, GPK, CP, JB, **Epidemiologic Data Management and Analysis:** DWC, MJD, IWP, AJ, GCF, BSY, SG, DB, HS, **Genomic Data Management and Analysis:** GKM, IWP, KMB, TCF, DHO, **Laboratory Technical Assistance:** CL, ACB, **Supervision:** RPW, JET, TCF, HLK, DHO, MEK.

## Abstract

University settings have demonstrated potential for COVID-19 outbreaks, as they can combine congregate living, substantial social activity, and a young population predisposed to mild illness. Using genomic and epidemiologic data, we describe a COVID-19 outbreak at the University of Wisconsin (UW)–Madison. During August – October 2020, 3,485 students tested positive, including 856/6,162 students living in residence halls. Case counts began rising during move-in week for on-campus students (August 25-31, 2020), then rose rapidly during September 1-11, 2020. UW-Madison initiated multiple prevention efforts, including quarantining two residence halls; a subsequent decline in cases was observed. Genomic surveillance of cases from Dane County, where UW-Madison is located, did not find evidence of transmission from a large cluster of cases in the two residence halls quarantined during the outbreak. Coordinated implementation of prevention measures can effectively reduce SARS-CoV-2 spread in university settings and may limit spillover to the community surrounding the university.

## Introduction

SARS-CoV-2, the virus that causes the coronavirus disease 2019 (COVID-19), can spread rapidly within congregate settings, including institutions of higher education (IHEs).^1,2^ During the fall of 2020, as IHEs around the United States resumed in-person instruction, IHE-associated SARS-CoV-2 cases began to rise.^3^ By February 2021, more than 530,000 COVID-19 cases linked to American IHEs had been identified.^4^ In many IHE settings that are populated significantly by young adults ages 18-24,^5^ there is less susceptibility to severe COVID-19 disease as compared to older populations (65+).^6^ Adhering to physical distancing is also particularly challenging for young people, for whom interaction with peers and social networks is important.^7^

As students returned to in-person learning, high-density clustering with on-campus housing and recreation potentially increased transmission and may have resulted in community outbreaks.^8-12^ Whole genome sequencing (WGS) provides an enhanced method for tracing cases. WGS relies on patterns of genome variation that eventually compose viral lineages, to reveal links between individuals that might not be apparent by case counts alone. This genome variation serves as a fingerprint, enabling tracking of specific SARS-CoV-2 lineages through space and time.^13-19^ One study using whole genome sequencing data suggested that SARS-CoV-2 transmission chains beginning or proliferating on IHE campuses may lead to spread within the community where the IHE is located, including to populations at higher risk of severe disease.^11^ Therefore, enhanced strategies to identify and prevent SARS-CoV-2 spread on IHE campuses and between IHEs and the community are needed.

Here, we use epidemiologic and genomic data to describe an outbreak of SARS-CoV-2 infection at the University of Wisconsin – Madison (UW-Madison) shortly after reopening for the Fall 2020 semester. We report the trajectory of the outbreak, including cumulative incidence within residence halls and attack rates among roommates, and describe measures taken to reduce transmission. In addition, using genomic data, we investigate whether UW-Madison-associated SARS-CoV-2 lineages may have spread into the community surrounding UW-Madison and discuss broader implications for IHE in preventing and responding to COVID-19 outbreaks.

## Methods

### Setting

UW-Madison is a large public university in the Midwest region of the United States, with approximately 45,540 enrolled students and 23,917 staff during the Fall of 2020.^20^ UW-Madison offered a combination of in-person and virtual classes for the Fall 2020 semester. Undergraduate students living in on-campus residence halls moved in on pre-assigned days during August 25-31, 2020. They were tested for SARS-CoV-2 on move-in day and were subsequently required to undergo a screening test every two weeks regardless of symptoms. Appointment-based testing for all students and staff was also made available free of charge. All specimens were collected via anterior nasal swabs, and testing was conducted using FDA-authorized real-time reverse transcription polymerase chain reaction (rRT-PCR) tests. At the beginning of the semester, UW-Madison also instituted a mandatory COVID-19 student pledge, which required mask usage at all times (except within students’ own rooms), physical distancing when possible, self-monitoring for symptoms, and limited gatherings in accordance with local public health guidelines.^21^ Students were provided a symptom screening tool to facilitate symptom self-monitoring; those screening positive were instructed to schedule a test and self-isolate (except to receive medical care) until a negative test result was received.

Isolation facilities were established in designated residence halls to temporarily house students living on-campus who had tested positive for SARS-CoV-2. Quarantine facilities were established in local hotels to temporarily house students living on-campus who had been in close contact with a person who tested positive for SARS-CoV-2. Roommates of students living in residence halls that tested positive were automatically moved into quarantine facilities, while the UW-Madison case investigation team elicited additional residence hall contacts who were then quarantined. Students identified as close contacts (defined as being within 6 feet of an in infected person for at least 15 minutes within a 24-hour period from 2 days before illness onset or positive specimen collection through the end of isolation) were quarantined in individual single rooms in hotels for 14 days, with meals delivered to the rooms, and tested for SARS-CoV-2 during the first and second week of quarantine. If a quarantined student tested positive, they isolated within the same location that they were quarantining in. Students testing positive that had not been in quarantine were transferred to designated isolation residence halls on campus. Isolation lasted for 10 days after symptom onset for those who were symptomatic, or 10 days after positive specimen collection for those who were asymptomatic, consistent with Centers for Disease Control and Prevention (CDC) recommendations.^22^

As the semester progressed, some modifications to the quarantine procedure were required due to the large volume of cases, particularly in two residence halls on campus (Residence Halls A and B). Given the high frequency of positivity within these two halls during universal testing events, all student residents of Residence Halls A and B were asked to quarantine within their residence hall for two weeks. This decision was made to mitigate transmission within Residence Halls A and Band prevent spread to other members of the campus and local community. During the quarantine period in these two residence halls, students were asked to wear a face covering when leaving their room, refrain from congregating, self-monitor for symptoms, test onsite, and stay in their residence hall. During the residence hall quarantines, residents testing positive were moved to an isolation facility, while roommates of residents testing positive initially quarantined within their residence hall room. Roommates of residents testing positive were not initially moved to an alternative quarantine facility during the full-hall quarantines as they were being frequently tested within the residence halls, and there were space and time limitations involved in moving roommates into alternate quarantine facilities. However, approximately one week into the residence hall quarantine, roommates of positive cases were moved to alternative quarantine facilities, student resident’s personal quarantine based on close contact with their roommate would end after the full-hall quarantine. Students could quarantine at their (permanent) home rather than in the residence hall if they chose; they could then return to the residence hall after the quarantine ended after providing a negative test result. Food was provided during designated time periods for residents of these two residence halls. Resident advisors checked in on students virtually, and mental health resources were provided by UW-Madison.

In addition to UW-Madison-specific mitigation measures outlined above, county-level ordinances passed earlier in the summer also applied to the UW-Madison community. As of July 13, 2020, Dane County Emergency Order #8 mandated the use of face coverings when in public, limited the size of public gatherings (10 people indoors, 25 people outdoors), limited restaurant capacity to 25%, and closed bars except for (a) ordering/pick-up and payment of food or beverage for takeout, or (b) providing outdoor seating with at least six feet distance between customers who were not members of the same household.^23^

### Epidemiologic data analysis

Wisconsin Department of Health Services public health surveillance system data (i.e., Wisconsin Electronic Disease Surveillance System, or WEDSS) were used to describe demographic characteristics, the location of on-campus clusters, and symptoms of COVID-19 cases. A UW-Madison-affiliated SARS-CoV-2 infection was defined as a positive SARS-CoV-2 rRT-PCR test result in a specimen collected from a UW-Madison student or staff member during August 1 – October 31, 2020. Daily percent positivity (positive SARS-CoV-2 specimens collected on a given day divided by the total number of specimens collected) and attack rates within residence halls were calculated using data collected by the campus testing program. Campus testing data were merged with University Housing data, which includes the residence hall and room number of each student living in on-campus housing, to determine specific housing location for students living on-campus as of September 22, 2020. A total of 19 residence halls with student populations ranging from 26-1195 individuals were included in the analysis. To calculate attack rates among roommates of positive cases, index cases were defined as the resident with the first positive SARS-CoV-2 test result within a room in a residence hall. Susceptible students were defined as residents sharing a room with an index case that had not previously tested positive for SARS-CoV-2. Roommate attack rates were defined as the proportion of susceptible students who had a positive SARS-CoV-2 specimen collected within 2-14 days after the index case specimen collection, consistent with the virus incubation period.^24^

Epidemiologic data analyses were performed using SAS software, version 9.4 (SAS Institute, Cary, NC), and RStudio, version 1.2.1335 (RStudio Team, Boston, MA).

### Whole Genome Sequencing

#### Sample selection criteria

Sequences for this study were derived from 262 anterior nasal swab samples from UW-Madison students living in the two residence halls experiencing the largest outbreaks, Residence Halls A and B, with collection dates between September 8-22, 2020. *Viral RNA (vRNA) isolation*

Anterior nasal swabs received in viral transport medium (VTM) were centrifuged at 21,130 x g for 30 seconds at room temperature. Viral RNA (vRNA) was extracted from 100 μl of VTM using the Viral Total Nucleic Acid Purification kit (Promega, Madison, WI, USA) on a Maxwell RSC 48 (Promega, Madison, WI, USA) instrument and was eluted in 50 μL of nuclease free H_2_O. All genomic samples were processed using a modified ARTIC tiled amplicon approach.^25,26^

#### Complementary DNA (cDNA) generation

cDNA was synthesized using a modified ARTIC Network approach.^25,27^ Briefly, vRNA was reverse transcribed with SuperScript IV Reverse Transcriptase (SSIV RT) (Invitrogen, Carlsbad, CA, USA) using random hexamers and deoxynucleotide triphosphates (dNTPs). Reaction conditions were as follows: 1 μL of random hexamers and 1 µL of dNTPs were added to 11 μL of sample RNA, heated to 65°C for 5 minutes, then cooled to 4°C for 1 minute. Final concentrations were 2.5µM for random hexamers and 0.5µM for dNTPs. Then 7 μL of a master mix (4 μL 5x RT buffer, 1 μL 0.1M dithiothreitol (DTT), 1 µL RNaseOUT RNase Inhibitor, and 1 μL SSIV RT) were added and incubated at 42°C for 10 minutes, 70°C for 10 minutes, and then 4°C for 1 minute.

#### Multiplex PCR to generate SARS-CoV-2 genomes

A SARS-CoV-2-specific multiplex PCR for Nanopore sequencing was performed, similar to amplicon-based approaches as previously described.^25,27^ In short, primers for 96 overlapping amplicons spanning the entire genome with amplicon lengths of 500 bp and overlapping by 75 to 100 bp between the different amplicons were used to generate cDNA. Primers used in this manuscript were designed by ARTIC Network and are shown in supplementary table 1. cDNA (2.5□μL) was amplified in two multiplexed PCR reactions using Q5 Hot-Start DNA High-fidelity Polymerase (New England Biolabs, Ipswich, MA, USA) using the following cycling conditions: 98°C for 30 seconds, followed by 25 cycles of 98°C for 15 seconds and 65°C for 5 minutes, followed by an indefinite hold at 4°C. Following amplification, samples were pooled before Oxford Nanopore Technologies (ONT) library prep.

**Table 1:**
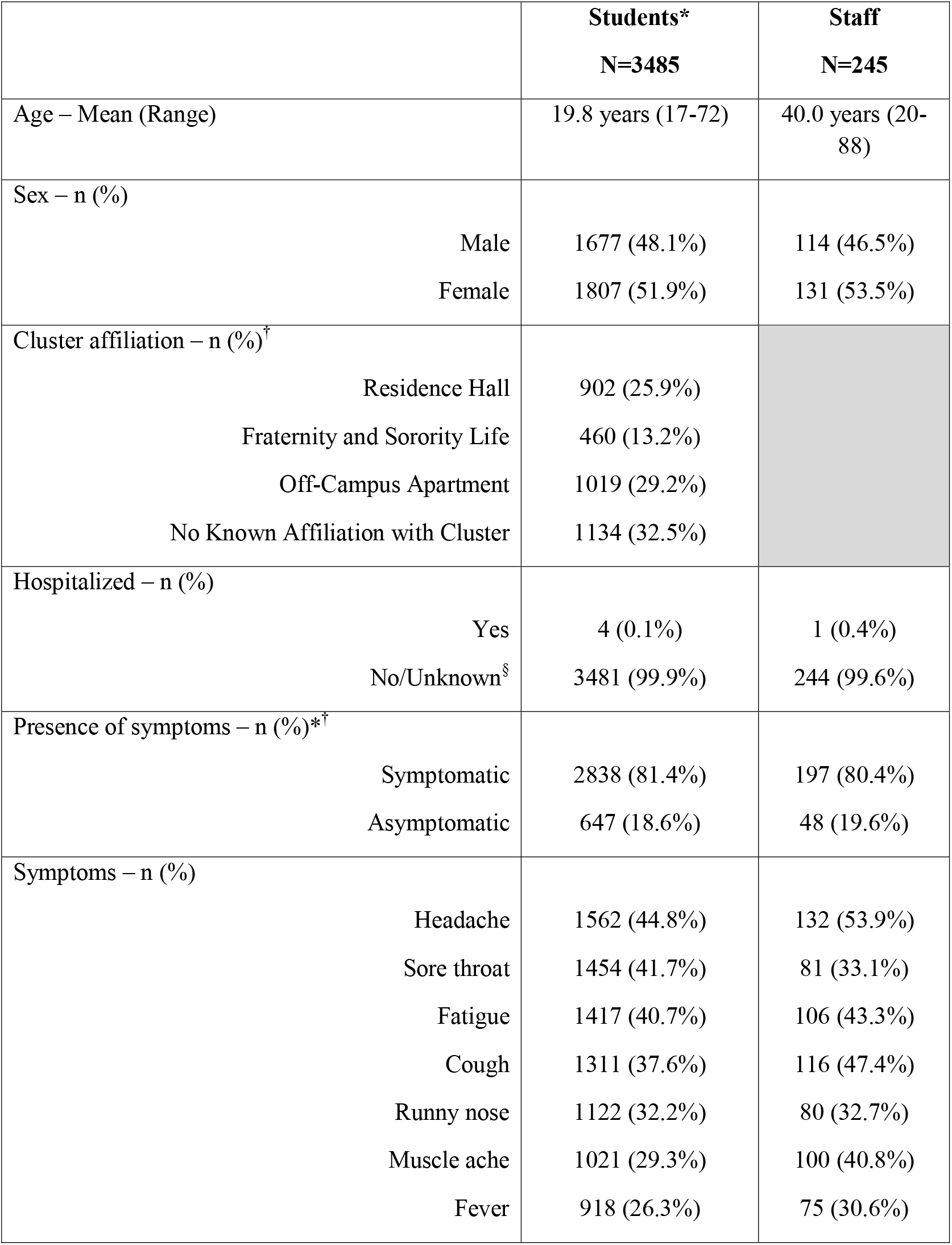
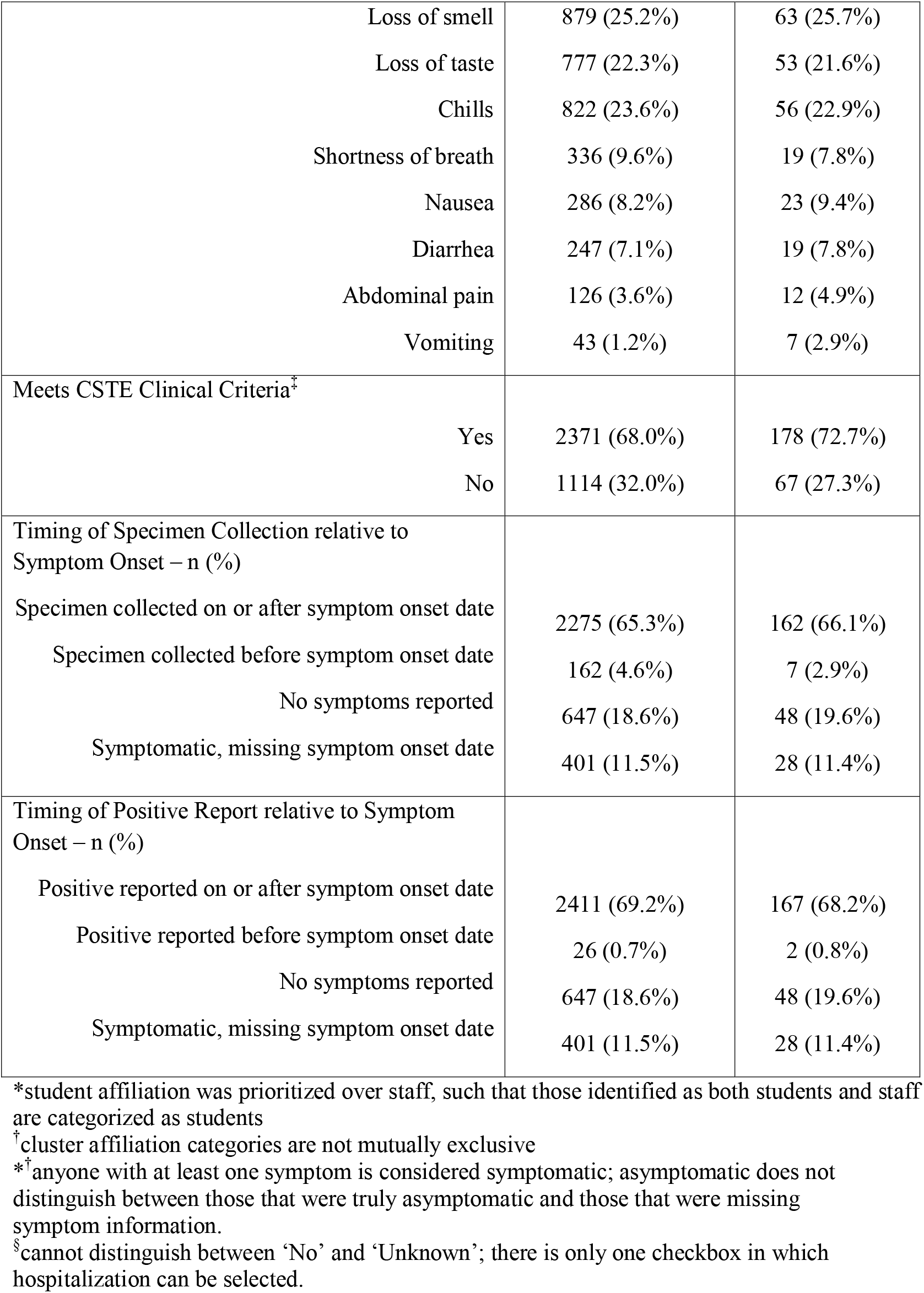
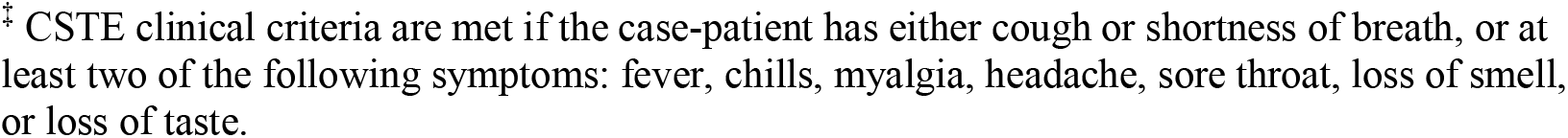
Characteristics of University of Wisconsin-Madison Student and Staff COVID-19 Cases, Dane County, Wisconsin, August 1 - October 31, 2020.

|  | **Students\***<br>**N=3485** | **Staff**<br>**N=245** |
| --- | --- | --- |
| Age – Mean (Range) | 19.8 years (17-72) | 40.0 years (20-88) |
| Sex – n (%) |  |  |
| Male | 1677 (48.1%) | 114 (46.5%) |
| Female | 1807 (51.9%) | 131 (53.5%) |
| Cluster affiliation – n (%)<sup>†</sup> |  |  |
| Residence Hall | 902 (25.9%) |  |
| Fraternity and Sorority Life | 460 (13.2%) |  |
| Off-Campus Apartment | 1019 (29.2%) |  |
| No Known Affiliation with Cluster | 1134 (32.5%) |  |
| Hospitalized – n (%) |  |  |
| Yes | 4 (0.1%) | 1 (0.4%) |
| No/Unknown<sup>§</sup> | 3481 (99.9%) | 244 (99.6%) |
| Presence of symptoms – n (%)<sup>*†</sup> |  |  |
| Symptomatic | 2838 (81.4%) | 197 (80.4%) |
| Asymptomatic | 647 (18.6%) | 48 (19.6%) |
| Symptoms – n (%) |  |  |
| Headache | 1562 (44.8%) | 132 (53.9%) |
| Sore throat | 1454 (41.7%) | 81 (33.1%) |
| Fatigue | 1417 (40.7%) | 106 (43.3%) |
| Cough | 1311 (37.6%) | 116 (47.4%) |
| Runny nose | 1122 (32.2%) | 80 (32.7%) |
| Muscle ache | 1021 (29.3%) | 100 (40.8%) |
| Fever | 918 (26.3%) | 75 (30.6%) |
| Loss of smell | 879 (25.2%) | 63 (25.7%) |
| Loss of taste | 777 (22.3%) | 53 (21.6%) |
| Chills | 822 (23.6%) | 56 (22.9%) |
| Shortness of breath | 336 (9.6%) | 19 (7.8%) |
| Nausea | 286 (8.2%) | 23 (9.4%) |
| Diarrhea | 247 (7.1%) | 19 (7.8%) |
| Abdominal pain | 126 (3.6%) | 12 (4.9%) |
| Vomiting | 43 (1.2%) | 7 (2.9%) |
| Meets CSTE Clinical Criteria<sup>‡</sup> |  |  |
| Yes | 2371 (68.0%) | 178 (72.7%) |
| No | 1114 (32.0%) | 67 (27.3%) |
| Timing of Specimen Collection relative to Symptom Onset – n (%) |  |  |
| Specimen collected on or after symptom onset date | 2275 (65.3%) | 162 (66.1%) |
| Specimen collected before symptom onset date | 162 (4.6%) | 7 (2.9%) |
| No symptoms reported | 647 (18.6%) | 48 (19.6%) |
| Symptomatic, missing symptom onset date | 401 (11.5%) | 28 (11.4%) |
| Timing of Positive Report relative to Symptom Onset – n (%) |  |  |
| Positive reported on or after symptom onset date | 2411 (69.2%) | 167 (68.2%) |
| Positive reported before symptom onset date | 26 (0.7%) | 2 (0.8%) |
| No symptoms reported | 647 (18.6%) | 48 (19.6%) |
| Symptomatic, missing symptom onset date | 401 (11.5%) | 28 (11.4%) |

\*student affiliation was prioritized over staff, such that those identified as both students and staff are categorized as students
<sup>‡</sup>cluster affiliation categories are not mutually exclusive
\*<sup>†</sup>anyone with at least one symptom is considered symptomatic; asymptomatic does not distinguish between those that were truly asymptomatic and those that were missing symptom information.
<sup>§</sup>cannot distinguish between ‘No’ and ‘Unknown’; there is only one checkbox in which hospitalization can be selected.

#### Library preparation and sequencing

Amplified PCR product was purified using a 1:1 ratio of AMPure XP beads (Beckman Coulter, Brea, CA, USA) and eluted in 30 μL of water. PCR products were quantified using Qubit dsDNA high-sensitivity kit (Invitrogen, USA) and were diluted to a final concentration of 1 ng/μl. A total of 5 ng for each sample was then made compatible for deep sequencing using the one-pot native ligation protocol with Oxford Nanopore kit SQK-LSK109 and its Native Barcodes (EXP-NBD104 and EXP-NBD114). Specifically, samples were end-repaired using the NEBNext Ultra II End Repair/dA-Tailing Module (New England Biolabs, Ipswich, MA, USA). Samples were then barcoded using 2.5 µL of ONT Native Barcodes and the Ultra II End Repair Module. After barcoding, samples were pooled directly into a 1:1 concentration of AMPure XP beads (Beckman Coulter, Brea, CA, USA) and eluted in 30 µL of water. Samples were then tagged with ONT sequencing adaptors according to the modified one-pot ligation protocol. Up to 24 samples were pooled prior to being run on the appropriate flow cell (FLO-MIN106) using the 72 hr run script.

#### Processing raw ONT data

Sequencing data were processed using the ARTIC bioinformatics pipeline scaled up using on-campus computing cores (https://github.com/artic-network/artic-ncov2019). The entire ONT analysis pipeline is available at https://github.com/gagekmoreno/SARS-CoV-2-in-Southern-Wisconsin.

#### Phylogenetic analysis

All available full-length sequences from Dane County through January 31, 2021, were used for phylogenetic analysis using the tools implemented in Nextstrain custom builds (https://github.com/nextstrain/ncov).^28,29^ In addition to the 262 samples sequenced from students living in Residence Halls A and B, 875 samples were sequenced from individuals tested at University of Wisconsin Hospital and Clinics (UWHC) from September 1, 2020 – January 31, 2021, representing approximately 3% of all cases within Dane County, where UW-Madison is located, during the time period. Those utilizing UWHC testing services included community members receiving pre-operative testing, employees, inpatient and emergency department patients, patients from associated hospitals, and those with known exposures. Of these 875 UWHC samples sequenced, 714 were collected on or after September 23, 2020, when the quarantine of Residence Halls A and B ended. This convenience sample of sequences was used to assess strains circulating within the Dane County community following the UW-Madison outbreak. Time-resolved and divergence phylogenetic trees were built using the standard Nextstrain tools and scripts. We used custom scripts written in Python to filter and clean metadata.

#### Analyses comparing roommate sequences

SARS-CoV-2 accumulates approximately one fixed mutation every other transmission event. ^30,31^ Therefore, if SARS-CoV-2 is directly transmitted from one individual to another, viruses from the two individuals in this “transmission pair” are expected to differ by ≤1 consensus single nucleotide variant (SNV). To test the hypothesis that roommates are more likely to have similar viral sequences than non-roommate pairs, we linked data from 33 roommate pairs in which both roommates had sequencing data and performed a permutation test comparing the percent overlap in single nucleotide polymorphism (SNP) identities between roommate pairs and random pairs of sequences derived from Residence Halls A and B. We performed a Mann-Whitney U test to compare the amount of diversity shared in roommate pairs and random pairs.

### Ethical approvals

A waiver of HIPAA Authorization was obtained by the Western Institutional Review Board (WIRB #1-1290953-1) to obtain the clinical specimens for whole genome sequencing. This analysis was reviewed by CDC and was conducted consistent with applicable federal law and CDC policy.^*^

These activities were determined to be non-research public health surveillance by the Institutional Review Board at UW-Madison.

## Results

### Demographics, symptom presentation, and measures to reduce transmission

During August 1 – October 31, 2020, 3,485 students and 245 staff affiliated with UW-Madison tested positive for SARS-CoV-2 by rRT-PCR, out of an overall enrollment of approximately 45,540 students and 23,917 staff **(Table 1)**. Cases in fraternity and sorority life (FSL) and other off-campus housing began rising before residence hall move-in week. UW-associated cases peaked during the week of September 6-12, 2020; soon after, cases began declining, with a sustained decline through September and consistently low case counts in October (**Figure 1**). Most student (81.4%) and staff (80.4%) case-patients reported at least one symptom of COVID-19; 68.0% of students and 72.7% of staff met the Council of State and Territorial Epidemiologists clinical criteria for a COVID-19 case (**Table 1**).^32^ Hospitalization was rare for both students and staff (<1.0%). Specimen collection occurred before symptom onset for 4.6% of student cases, while a positive result was reported before symptom onset for 0.7% of student cases. Among student cases, 902 (25.9%) were associated with an on-campus residence hall, 1,019 (29.2%) were associated with off-campus housing clusters, and 460 (13.2%) were associated with FSL (**Table 1**); the remainder were not linked to housing-specific clusters.

**Figure 1:**
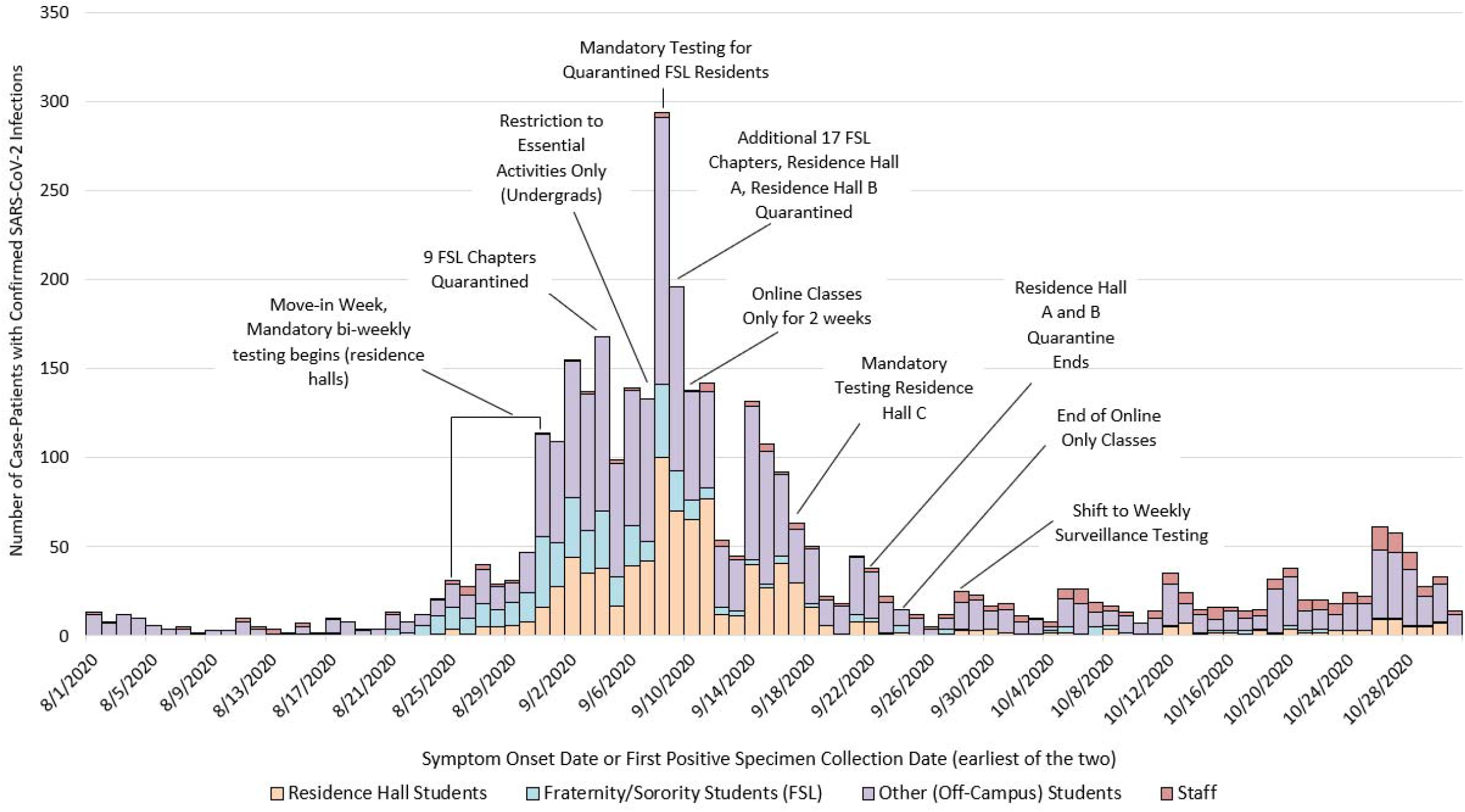
Overall Epidemic Curves of COVID-19 Cases among University of Wisconsin-Madison Students and Staff, Dane County, Wisconsin, August 1 – October 31, 2020*. *10 student case-patients affiliated with both a residence hall and a fraternity or sorority are categorized as Residence Hall Students. Student was considered the primary affiliation, such that any student who was also a staff member was categorized as a student.

Multiple mitigation measures were put into place to reduce transmission during the week of September 6-12, 2020. These included suspending in-person classes and other events, prohibiting non-sanctioned social activities, holding additional mass testing events, quarantining all students in Residence Halls A and B during September 9-23, 2020, as these residence halls were experiencing the highest percent positivity, and transitioning from biweekly to weekly COVID-19 screening tests for all students residing in any residence hall (**Figure 1**). The local health department, which maintains jurisdiction for off-campus housing, also required testing and quarantine for 26 FSL house chapters.

### Infections among students in residence halls

Across all residence halls, 5,820 of 6,162 students (94.4%) were tested during move-in week (August 25-31, 2020), with a mean turnaround time from test to result of two days (Interquartile Range: 1-2 days). Thirty-four students (0.6%) tested positive at move-in without documentation of a previous positive test in the last 90 days; these students were moved into isolation dorms. Overall, 856/6,162 (13.9%) students living in the 19 on-campus residence halls had a positive SARS-CoV-2 specimen collected through campus testing during August 25-October 31, 2020; attack rates in residence halls ranged from 1.9% to 31.9% (**Table 2)** during this time. Fifteen residence halls had attack rates of less than 10.0%, two had attack rates between 10.0% and 20.0%, and two had attack rates of over 20.0%. Residence Halls A and B accounted for 68.5% of all residence hall cases (586/856), but only 34.4% of all students living in residence halls (2,119/6,162) **(Figure 2)**; all students living in these two residence halls were asked to quarantine in place for 14 days starting September 9, 2020.

**Table 2:**
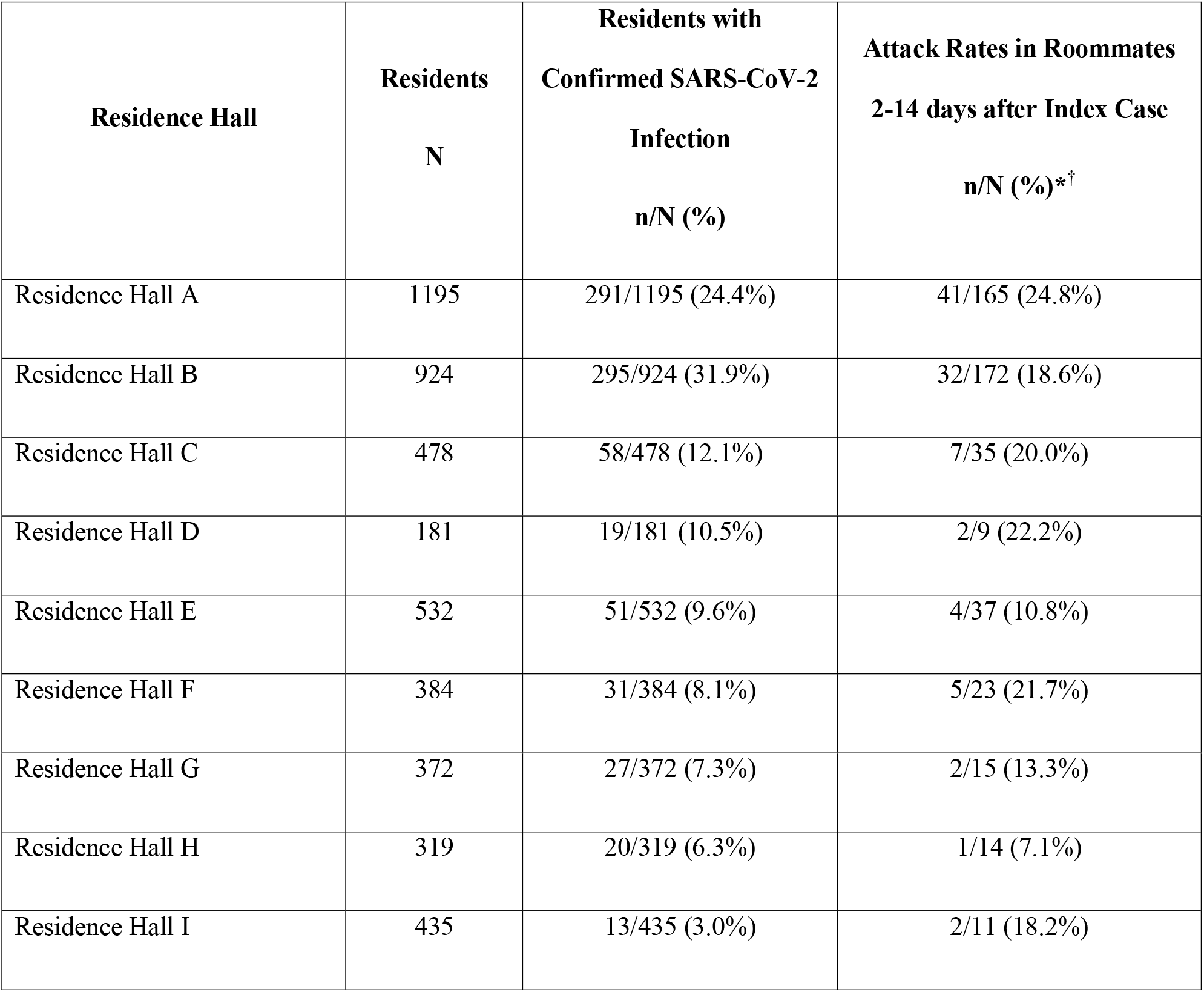
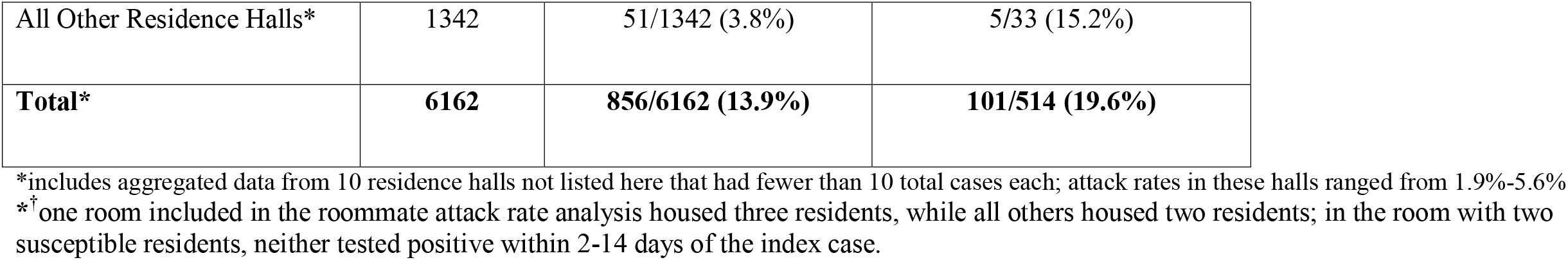
Attack Rates within Residence Halls and within Roommates for Residence Halls with ≥ 10 Cases, University of Wisconsin-Madison, Dane County, Wisconsin, August 25-October 31, 2020.

**Figure 2:**
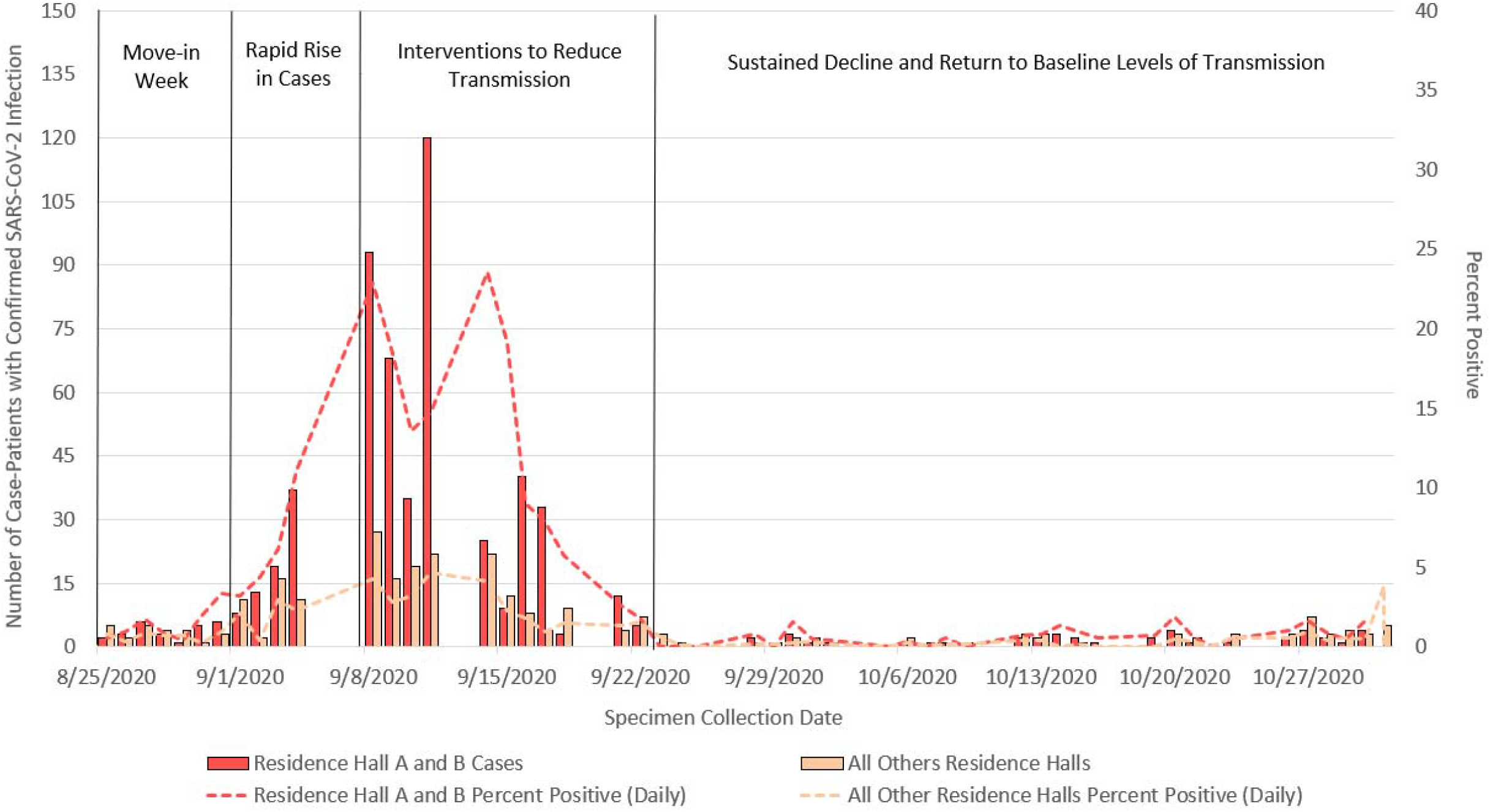
COVID-19 Epidemic Curves and Percent Positivity, University of Wisconsin-Madison Students Living in Residence Halls A and B vs. All Other Residence Halls, Dane County, Wisconsin, August 25 – October 31, 2020

In addition, we used a divergence phylogeny, colored by residence hall, to compare the number of mutations present in each sequence relative to the initial SARS-CoV-2 reference (Genbank: MN908947.3). If Residence Halls A and B had distinct, but contemporaneous outbreaks, we might expect viral sequences from the two halls to segregate into distinct taxa on a divergence tree. However, **Figure 3c** illustrates that substantial mixing of viral genetic lineages between the two residence halls occurred, indicating that outbreaks of COVID-19 within these residence halls were not distinct and resulted from intermingling between residents.

**Figure 3:**
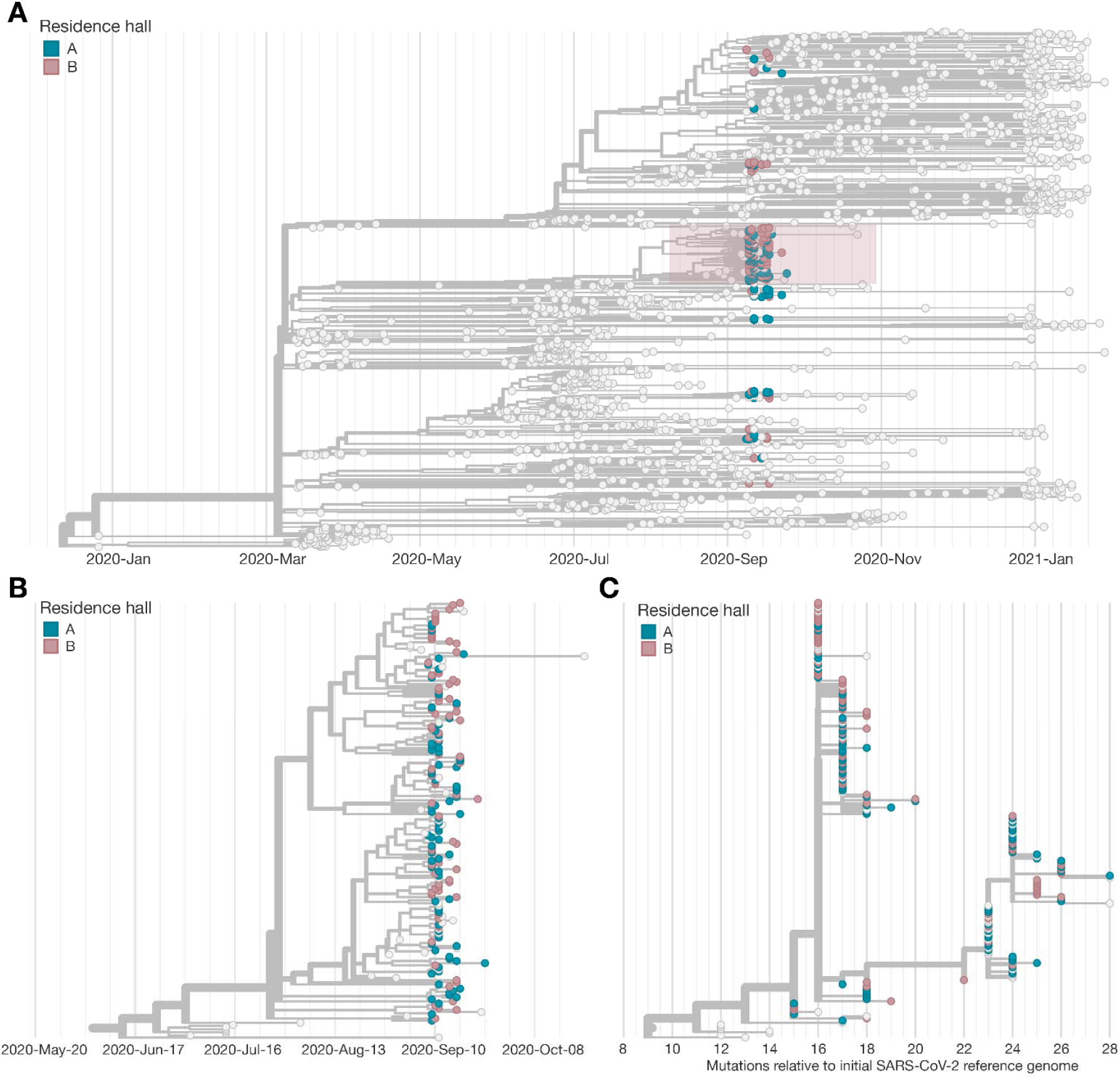
Phylogenetic Tree of the SARS-CoV-2 Outbreak in Residence Halls A and B, University of Wisconsin-Madison, Dane County, Wisconsin, January 2020 – January 2021. (a) Phylogenetic tree of all cases sequenced in Dane County, Wisconsin (light grey tips) during January 2020 – January 2021; sequencing of case-patients living on campus in Residence Halls A and B in blue and pink, respectively. (b) Zoomed in phylogenetic tree of the large cluster of cases associated with Residence Halls A and B (shown in the red box in Figure 3a) during the September 2020 outbreak. (c) Number of mutations relative to the initially identified SARS-CoV-2 genome in Wuhan (Genbank: MN908947.3) during the outbreak in Residence Halls A and B.

### Whole genome sequencing among students from two residence halls

We sequenced complete viral genomes from 262 (44.7%) of the 586 specimens taken from students living in Residence Halls A and B (**Figure 3**). Using a Dane County-centric phylogeny, we visualized the relationship of SARS-CoV-2 viruses circulating in Residence Halls A and B (**Figure 3**). Almost two-thirds of sequences from the residence halls (172/262; 65.6%) form a cluster in the 20A clade (PANGO lineage B.1.369) (**Figure 3b**). This cluster contains a unique spike mutation encoding a glutamic acid-to-glutamine substitution at spike residue 780 (S E780Q), which was not seen in Dane County prior to this outbreak. This mutation was not subsequently found in 467 sequenced specimens from Dane County (out of 15,740 total positive tests, for a sequencing coverage of 2.96%) during November 11, 2020 -January 31, 2021.

The remaining 90 residence hall sequences clustered with 20A (32/262), 20G (30/262), 20C (24/262), and 20B (4/262) clades. Sequences clustering in these remaining clades were more closely related to viral lineages concurrently circulating in Dane County. Between September 23, 2020, and January 31, 2021, 75.3% (538/714) of new sequences in Dane County were classified as 20G, 15.1% (108/714) classified as 20A, 7.0% (50/714) classified as 20C, and 2.5% (18/714) classified as 20B. **Figure 4**, in which colors tips on the phylogenetic tree by the age of the individual providing the sample, shows the large cluster in Residence Halls A and B was almost exclusively among case-patients aged 17-23 years..

**Figure 4:**
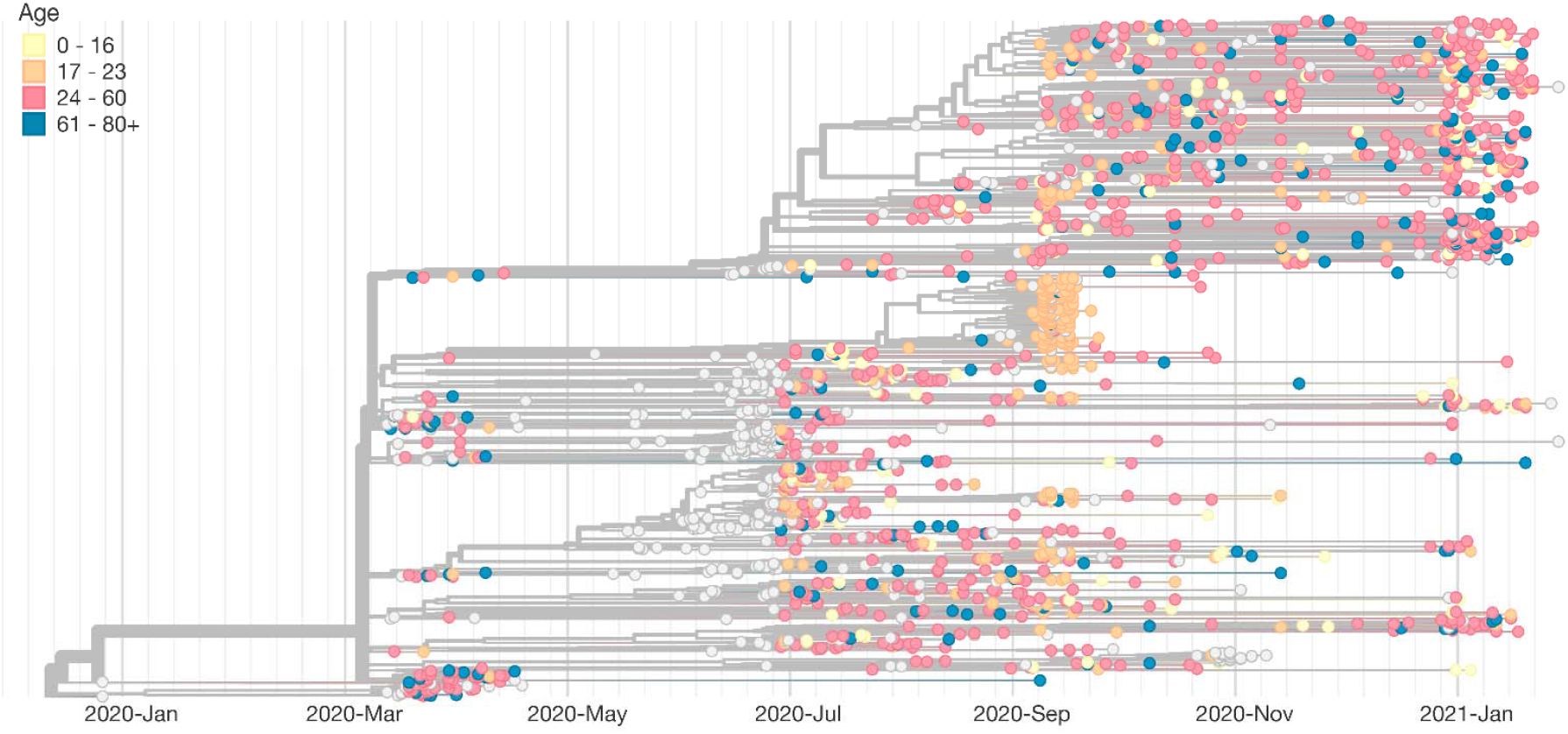
Phylogenetic Tree of the SARS-CoV-2 Specimens Sequenced in Dane County, Wisconsin, Coded by Age of Case-Patient Providing Specimen, January 2020 – January 2021

### Risk of transmission between roommates

Across all residence halls, 81.6% of residents had a roommate. Percent positivity was higher overall among students with roommates (15.4%) than those without roommates (7.3%) (p<0.0001). Of the 514 students who had a roommate test positive but had not yet tested positive themselves, 101 (19.6%) tested positive within 2-14 days. (**Table 2**).

Genetic distance comparisons between roommate pairs and non-roommate pairs within Residence Halls A and B revealed significantly higher levels of overlap in SNV identities between roommate pairs compared to random pairs. Specifically, 32/33 (97.0%) roommate pairs had viruses that contained 100.0% identical consensus sequences, while identical consensus sequences were found in only 1,062/33,930 (3.1%) of randomly assigned pairs (p<0.0001).

## Discussion

An outbreak of COVID-19 occurred at UW-Madison at the beginning of the fall semester despite nearly all students living in residence halls being tested when they moved in. Over the course of the investigation, almost 14.0% of students living in residence halls tested positive; those living with roommates were more likely to test positive. Shortly after the UW-Madison outbreak began, mitigation measures were rapidly implemented, and a rapid decline in cases was observed. Additionally, we did not detect evidence of transmission of the predominant viral lineages associated with Residence Halls A or B beyond these residence halls within Dane County. This suggests these interventions likely succeeded in preventing subsequent transmission and further spread into the Dane County community.

Testing at the time students moved into residence halls identified some introductions of SARS-CoV-2 onto campus, and UW-Madison subsequently facilitated isolation of infected students. However, the average two-day lag in turnaround time of test results meant that transmission might have occurred while these students were awaiting their test results. Students were only asked to self-quarantine if they had symptoms, but not specifically while awaiting test results. The on-campus residential move-in period represents a particularly high-risk situation in which students from different areas of the country (and world) with different exposure risks congregate together. Therefore, when implementing move-in testing, quarantining students until results have been received may help prevent transmission among asymptomatic students awaiting results.^33^ Move-in testing may also fail to identify students who have recently been infected and do not yet have detectable levels of SARS-CoV-2 virus.^34^ Move-in testing cannot prevent new infections from occurring after move-in if the virus is already circulating in the community and may have limited utility in an area of high transmission. Our results suggest that it is important to supplement move-in testing with ongoing serial testing and additional mitigation steps to effectively prevent ongoing transmission and community spread.

UW-Madison implemented serial screening testing every other week for students in residence halls with relatively short turnaround time (two days on average), enabling identification and isolation of those with symptomatic and asymptomatic SARS-CoV-2 infections, quarantine of roommates, and contact tracing to identify additional close contacts. Still, more frequent testing of students may have assisted in more rapid detection of cases and initiated isolation and quarantine procedures more quickly, preventing some of the spread observed in this investigation. A modeling study of COVID-19 spread within institutions of higher education (IHEs) suggested that frequent testing (e.g., every two days) would be needed to control the spread of the virus.^35^ In addition, while two-day turnaround time is relatively fast for PCR-based diagnostic testing, in a rapidly expanding outbreak in a congregate setting, this may still have allowed infected students to transmit SARS-CoV-2 before a positive result required them to isolate. Recognizing this potential for rapid spread, UW-Madison has modified their testing strategy for the Spring 2021 semester, increasing the frequency of testing to twice per week for students living on-campus and living off-campus in nearby zip codes and has reduced turnaround time for results to less than 24 hours.^36,37^ Further evaluation of serial testing strategies, including UW-Madison’s modified strategy for the spring 2021 semester, is needed to determine optimal testing frequency in IHE settings and to prioritize populations for testing when capacity is limited. The high proportion of infected students who were symptomatic (>80.0%) suggests that even in young adults, SARS-CoV-2 infection is frequently associated with at least mild symptoms, reinforcing the importance of educating students on COVID-19 symptoms, symptom monitoring, testing, and self-isolation when symptoms develop (even if only mild).^38^

Roommates live in close contact with each other, providing substantial opportunities for transmission.^39^ At UW-Madison, roommates were not required to wear masks within their rooms; requiring them to do so would be impractical and unenforceable. Roommates of confirmed case-patients within residence halls had an estimated attack rate of 19.6%, and a larger proportion of students with roommates tested positive over the investigation period than those without. Further, SARS-CoV-2 genomes collected from 33 roommate pairs found a high proportion of identical sequences, suggesting transmission occurred either within the roommate pair or based on a shared exposure. Given the elevated risk of infection associated with having a roommate, efforts to reduce the density of residence halls, including single-occupancy rooms when available, may reduce transmission.^1^

Two residence halls accounted for over two-thirds of all confirmed cases among students living in residence halls during the investigation period, despite the fact that these two halls accounted for only one-third of students living in on-campus housing. Students living in Residence Halls A and B were significantly more likely to visit bars in September 2020 than students living in other residence halls located further away from bars, which may have contributed to transmission.^40^ Transmission may have occurred within the residence halls, but it may have also occurred in other undetected settings (e.g., bars, private residences, fraternities or sororities) possibly more frequently visited by residents of Residence Halls A and B than by students living in other residence halls.^40,41^ The sequencing data strongly suggest that the clusters in Residence Halls A and B were not independent and were the result of intermingling, providing additional evidence that transmission may have occurred at common dining, study, and recreation areas that both halls share.

Viral genome sequencing is an important tool in understanding the transmission dynamics between UW-Madison students and the broader community.^11,13-19^ Our sequencing data covering 44.7% of student case-patients living in Residence Halls A and B, 7.5% of all student case-patients, and 3.0% of community samples from Dane County, suggest that the large cluster of UW-Madison cases associated with these residence halls gave rise to few descendent infections, with little evidence that viruses from this cluster subsequently circulated at high frequencies in the community. This suggests that the series of mitigation interventions put in place by the UW-Madison, including the quarantines of Residence Halls A and B and the suspension of most non-essential on-campus activity for two weeks, may have prevented substantial community transmission.

This analysis is subject to limitations. Full lists of off-campus students and staff and their COVID-19 testing histories were not available; therefore, attack rates could only be calculated for students living in on-campus residence halls. Data related to race, ethnicity, and other social determinants of health that may have impacted case counts and disease progression were not examined. Occupancy levels remained fluid throughout the semester, but available data used for residence hall census calculations represented a single point in time at the end of the outbreak, when occupancy was lower than at the start of the semester. UW-Madison’s rapid implementation of a series of interventions targeting different populations limits our ability to determine the effectiveness of each individual intervention. Specimens from students living in Residence Halls A and B were initially targeted for sequencing to understand transmission patterns within and across these housing units. Therefore, our sequencing results should not be generalized to the campus at large, as transmission events may have occurred after campus-related clusters outside of Residence Halls A and B that we did not characterize. In addition, sequencing of Dane County specimens in Nextstrain represented a small proportion of the total number infections within the county (about 3.0%) and were sampled non-randomly among clients of one large testing provider. Therefore, it is possible that descendant infections from UW-Madison clusters occurred in Dane County but were not captured in the convenience sample from the community. Finally, roommate attack rate calculations were limited in that campus testing data did not include information on symptom onset. Some roommate transmission events may have been erroneously excluded based on testing timeframes, resulting in underestimation of roommate transmission events. On the other hand, some transmission events attributed to roommates may have been due to a shared exposure outside of their room, resulting in overestimation of roommate transmission events.

This investigation described an outbreak where COVID-19 spread rapidly among university students at UW-Madison. Given the swift rise in cases, being able to quickly identify outbreaks and rapidly implement mitigation strategies via a coordinated university-wide response in collaboration with public health authorities are critical in halting transmission, both within the campus community and to the broader local community. Our findings suggest important strategies for IHEs and universities in preventing outbreaks include move-in testing with quarantine while awaiting results and frequent subsequent serial screening testing with rapid turnaround times, in addition to physical distancing, refraining from large gatherings, mask usage, and symptom screening and monitoring. Large-scale quarantines in congregate living situations (such as in dorms) and suspension of on-campus activities may be effective when experiencing large-scale outbreaks, if implemented rapidly and effectively. Given the elevated risk of transmission among roommate pairs, strategies to reduce the density of residence halls and utilize single occupancy rooms when available may reduce the spread of infection. This investigation also demonstrates the use of genomic surveillance to provide a more comprehensive understanding of transmission dynamics both in specific outbreak settings and in the general population; these tools can be used by universities and health departments to monitor spillover into the community and inform campus and community mitigation efforts.

## Data Availability

All sequencing data is available on www.gisaid.org. Scripts for sequence data analysis is available at https://github.com/gagekmoreno/SARS-CoV-2-at-UW_Madison.

## Acknowledgements

This research was performed using the computing resources and assistance of the University of Wisconsin (UW)-Madison Center for High Throughput Computing (CHTC) in the Department of Computer Sciences. The CHTC is supported by UW-Madison, the Advanced Computing Initiative, the Wisconsin Alumni Research Foundation, the Wisconsin Institutes for Discovery, and the National Science Foundation, and is an active member of the Open Science Grid, which is supported by the National Science Foundation and the U.S. Department of Energy’s Office of Science. G.K.M. is supported by an NLM training grant to the Computation and Informatics in Biology and Medicine Training Program (NLM 5T15LM007359). This work was funded in part by the U.S. Centers for Disease Control and Prevention Contract #75D30120C09870: Defining the Role of College Students in SARS-CoV-2 Spread in the Upper Midwest.

## Footnotes

* 45 C.F.R. part 46.102(l)(2), 21 C.F.R. part 56; 42 U.S.C. Sect. 241(d); 5 U.S.C. Sect. 552a; 44 U.S.C. Sect. 3501 et seq.

## Notes

### Clinical Trial

N/A.

### Author Declarations

A waiver of HIPAA Authorization was obtained by the Western Institutional Review Board (WIRB #1-1290953-1) to obtain the clinical specimens for whole genome sequencing. This analysis was reviewed by CDC and was conducted consistent with applicable federal law and CDC policy. These activities were determined to be non-research public health surveillance by the Institutional Review Board at UW-Madison.

